# Blended care in patients with knee and/or hip osteoarthritis in physical therapy: a Delphi study on needs and preconditions

**DOI:** 10.1101/2022.10.25.22281495

**Authors:** F. Weber, C. Kloek, A. Arntz, C. Grüneberg, C. Veenhof

**Author notes:** Corresponding author: F. Weber, PT, M.Sc., Department of Applied Health Sciences, Division of Physiotherapy, Hochschule für Gesundheit (University of Applied Health Sciences), Gesundheitscampus 6-8, 44801 Bochum (Germany). Address all correspondence to Ms. Weber at. **Trial registration:** DRKS00023386. **Footnotes:** ^*^SoSci Survey GmbH, Munich, Germany. **List of declarations**. **Competing interests:** Nil. **Ethics approval:** The Ethics Committee of the University of Applied Health Sciences in Bochum approved this study. All participants gave written informed consent before data collection began.

## Abstract

**Introduction:** Osteoarthritis is a major public health concern. Despite existing evidence-based treatment options, the health care situation remains unsatisfactory. Digital care options, especially combined with in-person sessions seems to be promising. Therefore, the aim of this study was to investigate the needs, preconditions, barriers and facilitators of blended physical therapy.

**Methods:** This Delphi study consisted of interviews, an online survey and focus groups. Participants were physical therapists, patients with hip or knee osteoarthritis with or without experience in digital care and stakeholders of the health care system. In the first phase, interviews were conducted with patients and physical therapists. The interview guide was based on the “Consolidated Framework For Implementation Research”. The interviews focused on experiences with digital and blended care. Furthermore, needs, facilitators and barriers were discussed. In the second phase, an online survey and focus groups served the process to confirm the needs and collect preconditions.

**Results:** Nine physical therapists, seven patients and six stakeholders confirm that an increase of acceptance of digital care by physical therapists and patients is crucial. One of the most frequently mentioned facilitator was conducting regular in-person sessions. Physical therapists and patients conclude that blended physical therapy needs to be tailored to the patients’ needs. The reimbursement of blended physical therapy needs to be clarified.

**Discussion:** Most importantly, it is necessary to strengthen the acceptance of patients and physical therapists towards digital care. Overall, for development and implementation purposes, it is crucial to take the needs and preconditions into account.

## Introduction

Osteoarthritis (OA) is a major public health problem with a high prevalence worldwide, which will further increase in the next years due to the ageing population, rising obesity rate and people being physically inactive (1). In particular, the burden of OA on the health care system is expected to grow exponentially (1). While effective treatment is available, these conservative treatment options are still underutilized (2).

To facilitate the access to primary care and to reduce health-related costs, digital health care is a promising approach. Especially looking at the course of the COVID-19 pandemic, the potential of digital health has been demonstrated confirming that it is not just a megatrend (3). A general definition of digital health is the application of information and communication technologies across a broad range of activities, which are performed in health care (4). Combining in-person and digital health care is referred to as blended care (5). Linking the advantages of online and offline guidance and treatment yield positive outcomes (6, 7). Benefits of blended care include the stronger focus on patient empowerment and lower resource use compared to traditional in-person treatment (6, 8). In addition, blended care potentially increases and facilitates the patient’s self-management, for instance by supporting adherence to exercise recommendations (9). In the Netherlands, a blended physical therapy intervention, called *e-Exercise*, has already proven its potential for people with hip or knee OA (10). This *e-Exercise* intervention revealed the same effectiveness but less physical therapy sessions compared to traditional physical therapy (10).

However, it is important to note that blended care is not suitable in all cases, potentially because of preferences and motivation of patients, severity of illness, comorbidities, level of education, digital and health literacy (11, 12). In addition, it has to meet the needs of the physical therapists. Thus, to optimize the use and implementation of blended care approaches in an outpatient setting, it is important to involve both patients and physical therapists as well as other relevant stakeholders in the development process to take their needs and preconditions into account (13).

Therefore, the objective of this study is to get insight in the needs, preconditions, barriers and facilitators regarding blended physical therapy in patients with knee and hip OA from the perspective of patients, physical therapists and other stakeholders of the health care system.

## Methods

### Design

A Delphi method was used (14) aiming to get insight into needs, preconditions, facilitators and barriers with respect to the content, sequence and ratio of blended physical therapy. Established methodological criteria for reporting Delphi studies were followed to ensure quality (15). The study design is shown in Figure 1. This study was conducted in accordance with the Declaration of Helsinki (16). The ethics committee of the University of Applied Sciences Bochum approved the study (201116_Grüneberg, 04.01.2021).

**Figure 1.**
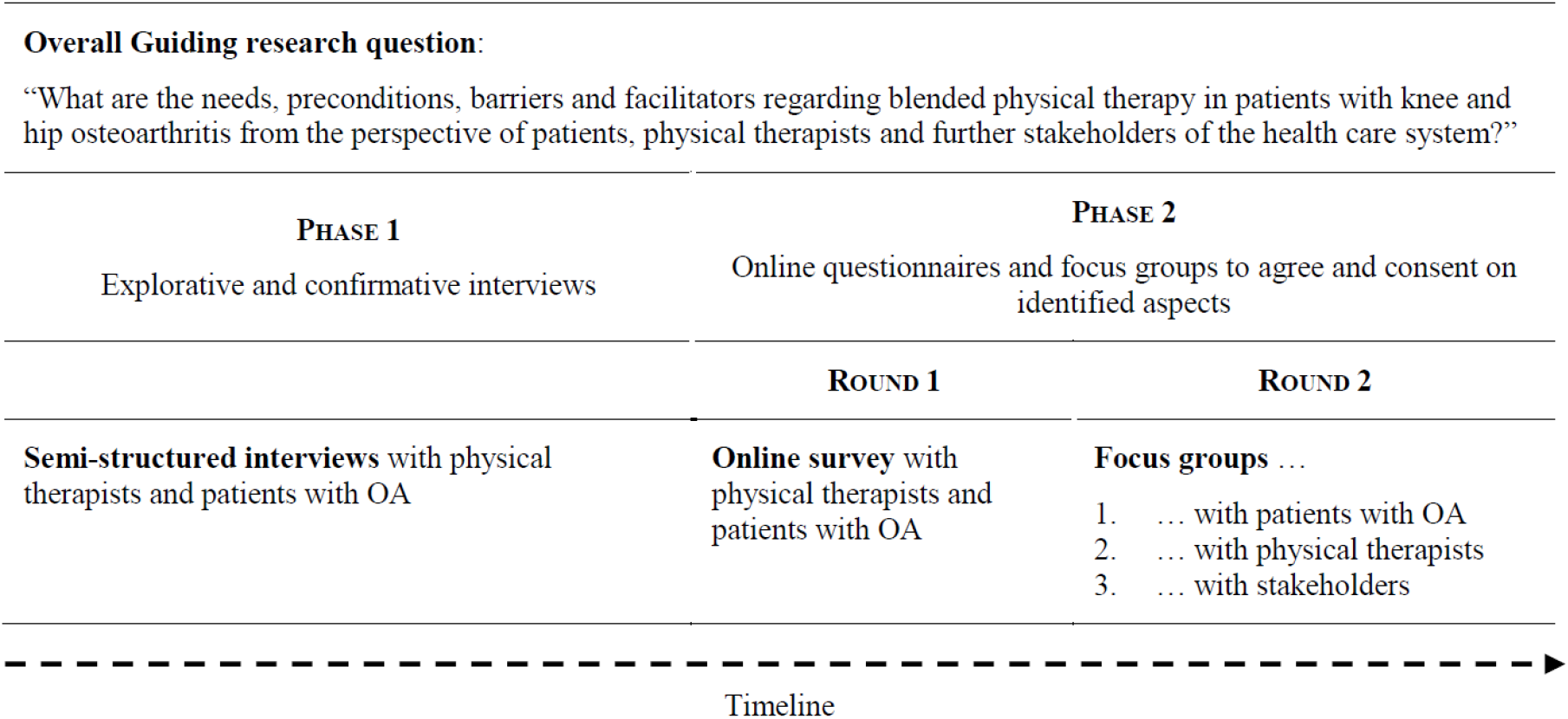
Study design and methodological description of the different phases of the Delphi process

### Participants

#### Physical therapists

We recruited physical therapists using the database of clinical cooperation partners of the University of Applied Sciences (Bochum) and through personal network. To be eligible, participants needed to have a (study) degree in physical therapy; work in an outpatient physical therapy setting; have at least five years of experience in treating patients with OA; give informed consent; be able to understand and speak German; have access to the internet and own a digital device.

#### Patients

Participating physical therapists were asked to contact eligible patients with OA and sent them an information letter regarding the study. Furthermore, patients were recruited through personal network, e.g. via patient associations. Inclusion criteria for the patients were a medically diagnosed idiopathic OA of the knee or the hip and a signed informed consent. Further criteria were to be able to understand and speak German; to have received at least one prescription for physical therapy regarding their OA related symptoms; to own a digital device and to have internet access.

The aim was to recruit both physical therapists and patients who already had experience with digital health care in any context, as well as physical therapists and patients who did not have this experience. Participants were recruited until saturation was reached, which was when no new information would be identified from the last two interviews (17).

#### Stakeholders of the health care system

In order to obtain a broad distribution of participants, we aimed to recruit a member of a patient association, an owner of a physiotherapeutic practice, a physician, a politician in the field of health care, a person of a health insurance company, a representative of a company developing digital devices and a member of a physical therapy association. We recruited them through patient associations, assisted by a German physical therapy association and through personal network. To be eligible, participants needed to have at least five years of professional experience in their field; internet access; own a digital device; give signed informed consent and have sufficient skills in German.

### Procedure

The Delphi process consists of two phases, phase two was separated in two rounds (Figure 1).

#### Phase 1: explorative and confirmative interviews

Phase 1 was an explorative phase with the aim to capture different perspectives. Both patients and physical therapists filled out questionnaires regarding demographic data (age, gender, educational level and experience with digital/blended care) and their (digital) health literacy (HLS-EU-Q16 and eHEALS) (18, 19). Further, they were asked to participate in individual semi-structured interviews via telephone. Topics for the interviews were developed on the basis of the *Consolidated Framework For Implementation Research (CFIR)* (Suppl. Material 1) (20). The CFIR consists of the following five domains: (1) characteristics of the individuals involved, (2) intervention characteristics, (3) inner setting, (4) outer setting and (5) the process of implementation (20). The process of implementation was not questioned, since there was no specific intervention to implement, yet. Each participant was asked about his/her experiences with digital health care, possible facilitators and barriers they experienced or would expect from digital and blended care in the four domains of the CFIR. In between, a short video (https://hs-gesundheit.sciebo.de/s/hiiSJDdS0zYNfIk) was presented during each interview, which showed an example of blended care (combination of in-person physical therapy, video conference and app) and gave a definition of blended care to create a common sense.

#### Phase 2: online questionnaire and focus groups to agree and consent on identified aspects

The aim of phase two, consisting of two rounds, was to get consensus about specific preconditions for blended care in physical therapy. The same group of physical therapists and patients were invited to fill out an (anonymous) online survey via a secured online platform^*^ in round one. Two researchers (AA and FW) translated the results of phase one into statements; the participants had to agree or to disagree on these statements (4-point Likert scale: “I completely disagree” (1) to “I completely agree” (4)). The online questionnaire was quantitatively evaluated and the results were used for round two of this phase. At the beginning of the second round, the results from the survey were briefly presented and the aim of the focus group was explained.

Three focus groups (patients, physical therapists and stakeholders) were conducted to agree and consent on results of the online questionnaires (Figure 1). In addition, the aim was to examine what essential preconditions are necessary to make blended physical therapy feasible in an outpatient practice.

### Data analysis

#### Phase 1

Two researchers (AA and FW) transcribed verbatim and coded the transcripts of the interviews. Data analysis of the interviews was performed based on the framework approach (21). Using explorative data analysis for each main topic from the interview scheme, citations were extracted and arranged into themes and subthemes. Subsequently, these themes were discussed between the researchers (AA, FW) until consensus was reached (Suppl. Material 2). Finally, all codes of each theme of every participant were displayed in a table (22). Next, one researcher (FW) examined the raw data again to ensure the robustness of the analytical process and to confirm that all data were indeed reflected in the coding. Transcription, coding, organization and analysis were done using MAXQDA Plus 2020, Windows Version 20.3.0.

#### Phase 2

Data from round one were exported from the secure online platform^*^ into an excel sheet. Demographics, data from the (digital) health literacy questionnaires, as well as data from the online survey was analyzed descriptively with SPSS (IBM SPSS Statistics 25). Results were analyzed by quantifying scores on each item from the survey and calculating percentages of patients and physical therapists, who chose a certain answer on the items.

In round two, focus groups were recorded in writing protocols. Data were categorized into the corresponding themes or subthemes of the interviews according to the CFIR domains. Categorization was discussed between two researchers (CG and FW) until consensus was reached. Data were screened regarding repetitions and each theme and corresponding subthemes were summarized.

## Results

Nine physical therapists and seven patients participated in the interviews and the online survey, five of the physical therapists and four of the patients took part in the focus groups, respectively. The third focus group consisted of six stakeholders and one physical therapist of the first phase.

### Characteristics of participants

For physical therapists of phase 1, saturation was reached after nine interviews. The characteristics of physical therapists are shown in Table 1.

**Table 1.** Characteristics of physical therapists

| Characteristics | Physical therapists (n = 9) |
| --- | --- |
| Age (*yr*), mean (SD) | 33.0 (6.5) |
| Sex, n (%) |  |
| male | 4 (44) |
| female | 5 (55) |
| Clinical experience (*yr*), mean (SD) | 9.4 (6.2) |
| Clinical experience in treating patients with OA (*yr*), mean (SD) | 9.4 (6.2) |
| Level of education, n (%) |  |
| Masters, diploma, state examination (university (of applied sciences)) | 1 (11)<br>6 (67) |
| Bachelors (university or university of applied sciences) | 2 (22) |
| Apprenticeship |  |
| Prior experience in online therapy, n (%) | 4 (44) |
| Working hours/ week (*hrs*), mean (SD) | 32.0 (12.5) |
| General health literacy (HLS-EU-Q16), n (%) |  |
| adequate | 4 (44) |
| problematic | 4 (44) |
| inadequate | 1 (11) |
| Digital health literacy (G-eHEALS), mean (SD) | 32 (7) |

<sup>a</sup>yr = year; SD = standard deviation; OA = osteoarthritis; hrs = hours; HLS-EU-Q16 = the European Health Literacy Survey Questionnaire (0 (low/no health literacy) to 16 high health literacy); G-eHEALS = German eHealth Literacy Scale (0-40 and higher score indicating better digital health literacy)

Concerning the patients in phase 1, saturation was achieved after seven interviews. Table 2 displays the characteristics of patients.

**Table 2.** Characteristics of patients

| Characteristics | Patients (n = 7) |
| --- | --- |
| Age (*yr*), mean (SD) | 59.9 (10.8) |
| Sex, n (%) |  |
| male | 4 (57) |
| female | 3 (43) |
| Osteoarthritis, n (%) |  |
| hip osteoarthritis | 3 (43) |
| knee osteoarthritis | 1 (14) |
| Both | 3 (43) |
| Time since diagnosis (*yr*), mean (SD) |  |
| hip osteoarthritis | 9.8 (8.3) |
| knee osteoarthritis | 7.0 (3.8) |
| Degree of self-reported limitations due to osteoarthritis, n (%) |  |
| fair | 2 (29) |
| mild | 5 (71) |
| Duration of physical therapy due to osteoarthritis-related symptoms (*yr*), mean (SD) | 3.6 (6.0) |
| Level of education, n (%) |  |
| high level of education | 5 (71) |
| low level of education | 2 (29) |
| Prior experience in online therapy, n (%) | 3 (43) |
| General health literacy (HLS-EU-Q16), n (%) |  |
| adequate | 7 (100) |
| Digital health literacy (G-eHEALS), mean (SD) | 32.6 (4.4) |

<sup>a</sup>yr = year; SD = standard deviation; OA = osteoarthritis; HLS-EU-Q16 = the European Health Literacy Survey Questionnaire (0 (low/no health literacy) to 16 high health literacy); G-eHEALS = German eHealth Literacy Scale (0-40 and higher score indicating better digital health literacy)

One physical therapist with experience in digital health joined the other stakeholders in the last focus group. The politician in the field of health care was not able to participate in the focus group.

The results in Table 3 summarize the needs and preconditions of the patients, physical therapists and the stakeholders regarding blended care, which are the final results of the two phases.

**Table 3.** Consensus of the needs and preconditions regarding blended physical therapy in the perspective of patients with osteoarthritis, physical therapists and stakeholders

|  |
| --- |
| **PERSONAL FACTORS (INDIVIDUAL)** |
| **Change of the role of physical therapists and gaining new competences**<br>*(Necessity of changing the role of physical therapists, patient and physical therapist being equal partners; new competences are necessary)* |
| **Attitudes and acceptance (Changing attitudes and increasing the acceptance for digital health)**<br>*(Necessity to change attitudes towards and acceptance for digital health)* |
| **INTERVENTION-RELATED FACTORS (BLENDED CARE)** |
| **Digital content and feature**<br>*(Educational components, information exchange and exercise program as digital content; important to include motivational strategies in the app, e.g. reminders; see Table 4)* |
| **Usability and operability**<br>*(Easy and intuitive app, necessity of user-friendliness, patient-friendly language, flexibility in decision-making, wide accessibility of the app, feedback through data)* |
| **Blended care concept (individualization, ratio and allocation)**<br>*(Individualization is necessary, integration of evidence-based information, regular face-to-face sessions, ratio of online and face-to-face 60:40 (online : face-to-face sessions), flexibility of online or face-to-face mode)* |
| **ORGANIZATIONAL FACTORS (INNER SETTING)** |
| **Practice setting (e.g. working conditions, personnel structures, hardware)**<br>*(Change of rooms, necessity of hardware, WLAN, software, positive influence on the working conditions, change of personnel structures, change of time schedules, necessity of interoperability of different programs)* |
| **Change of (interprofessional) cooperation/ communication**<br>*(Facilitation of interprofessional communication by online environment)* |
| **SYSTEM-RELATED FACTORS (OUTER SETTING)** |
| **Necessity to change efficiency (e.g. time and costs)**<br>*(Time to prepare, efficiency of time, increase in costs, who will pay?)* |
| **Necessity of clear structural conditions, e.g. rules regarding data protection and security**<br>*(Clear description of concept is necessary, prescription or integration in disease management program is necessary; legal basis; necessity of clear rules and legal aspects regarding data protection; data protection guidelines; implementation of advanced training/ skills)* |
| **Clear rules and roles before an implementation**<br>*(Development process of digital devices; responsibility for implementation process (stakeholders))* |

### Personal factors (Individual)

#### Change of the role of physical therapists and gaining new competences

A changing role of physical therapists was a central precondition for blended care, which received consensus of physical therapists and stakeholders. Different facets of changes have been mentioned; however, the main adjustment was seen in the patient-provider relationship. According to physical therapists, both should be on an equal level with the physical therapist being in a guiding role. There was a full consensus of the physical therapists that blended care has an essential impact to facilitate patient’s self-management and individual responsibility.

Patients also considered a healthy relationship with and trust in the physical therapist as a crucial precondition for blended care. Contrary to the perspective of physical therapists, passive interventions (and therefore in-person contact) were still one of the most important aspects of physical therapy for patients.

Patients and physical therapists considered adequate communication skills of both and a moderate level of health literacy of patients as necessary. In the perspective of physical therapists, a core competence of themselves within blended care was the need to be familiar with the technology used. All physical therapists and stakeholders concluded, that a further competence is the decision making, if the approach is useful and feasible for every patient. As a precondition for using digital health in physical therapy, they mentioned an adequate training of new competences for the physical therapists and gaining positive experiences with digital health for patients and physical therapists.

#### Attitudes and acceptance

All participants mentioned the COVID-19 pandemic as a facilitator for blended care, especially increasing the acceptance of digital health. Most of the physical therapists were open regarding digital health care. Patients needed and wanted to learn how to handle digital tools in advance. The acceptance of digital health care of patients varied, but in general, they recognized the convenience to exercise anytime and place and incorporating the therapy into their daily lives. Further preconditions to increase the acceptance of patients were the confidence in the physical therapist and sufficient time to learn and practice.

### Intervention-related factors (Blended care)

#### Digital content and feature

The vast majority of all participants considered educational components, information exchange and an exercise program as content, which can be carried out digitally. The results of the online questionnaire regarding the preferred mode of delivery are shown in Table 5 (Suppl. Material 3, Table 5).

All physical therapists agreed on the importance to integrate motivational strategies in the technology, e.g. activity trackers and reminders (Table 4).

**Table 4.** Preferred content and features of the online program within a blended physical therapy approach out of patient and physical therapist’s perspective

| **Content of the online program** | **Patients (n=7)** | **Physical therapists (n=9)** |
| --- | --- | --- |
| Exercise/ training plans, which include PA and exercises | 100% | 100% |
| Therapy/ treatment plans, which include goal appropriate exercises and treatment | 100% | 100% |
| Examination/ warning of red flags regarding the treatment of patients with OA | 86% | 100% |
| Test/ MI instructions, which are performed by the patient on his/her own or the physical therapist with the patient, e.g. 6MWT, TUG | 57% | 78% |
| Communication/ exchange with physicians or other professions | 57% | 56% |
| Information on relevant topics for patients with OA | 43% | 100% |
| Patient-reported outcome measures, e.g. KOOS, HOOS | 43% | 89% |

| Features of the online program | Patients (n=7) | Physical therapists (n=9) |
| --- | --- | --- |
| Chat for communication between physical therapists and patients | 86% | 78% |
| Documentation system for the physical therapist | 71% | 78% |
| Agenda with future physical therapy appointments | 71% | 33% |
| Video chat | 57% | 100% |
| Diary of patients to collect PA and exercises | 57% | 89% |
| Collecting/ capturing of data of the course of therapy of the patient | 57% | 78% |
| Reminder messages of appointments | 43% | 100% |
| Reminder messages of PA | 29% | 89% |

<sup>a</sup>PA = physical activity; OA = osteoarthritis; MI = measurement instrument; 6MWT = Six-Minute-Walking Test; TUG = Timed “Up & Go”-Test; KOOS = Knee Injury and Osteoarthritis Outcome Score; HOOS = Hip Disability and Osteoarthritis Outcome Score

Physical therapists perceived the digital program within blended care as a guiding tool, whereas patients saw digital components only as a supplement to regular in-person sessions. The results of the online survey including specific software features and content are presented in Table 4.

#### Usability and operability

Patients and physical therapists had the same opinion regarding the importance of technology being user-friendly. From their perspective, digital tools should be easy and intuitive to use.

#### Blended care concept

All participants agreed that blended care must be tailored to the patients’ individual needs. Participants considered in the online survey an average ratio of 60/40 (digital/in-person sessions) as optimal (Suppl. Material 3, Table 6). Physical therapists and patients consider that a first in-person session is crucial, the longer the treatment process, the less in-person sessions are necessary. Stakeholders stated that the needs of the patient, access to devices, state of condition and confidence in physical therapy, motivation of the patient, as well as a high level of patients’ self-management are factors, which influence the decision on the most appropriate therapy mode.

An academic education and several years of professional experience as a physical therapist were mentioned as preconditions, since it supports the decision on the therapy mode in the opinion of the stakeholders.

The stakeholders emphasized the value of “taking the physical therapist home”, which increases the sustainability of therapy in their point of view.

### Organizational factors (Inner setting)

#### Practice setting

Patients and physical therapists considered a separate room only for digital care (e.g. video conference) as necessary to be undisturbed, maintain privacy of the patient and having all equipment ready to use.

Physical therapists considered a change of practice structures as necessary. A proper time planning is important, e.g. to prepare online sessions. Table 7 summarizes the stated preconditions regarding a practice setting for the implementation of blended care (Suppl. Material 3, Table 7).

A precondition for blended care was that every user has access to digital devices and a stable internet connection. Physical therapists preferred tablets or laptops as hardware. Patients considered missing equipment and technical requirements as a barrier for blended care. They preferred a large screen on their digital devices. The stakeholders stated the importance of the interoperability of different systems, especially with already existing ones.

#### Change of (interprofessional) cooperation/ communication

The interviewed physical therapists expected a facilitation and simplification of the interprofessional communication and cooperation within blended care. For instance, data will be collected and stored in a more structured way and the treating physician would have the option to access the status or progress of the patient, in that way the communication between the physical therapist and physician is based on results and data. Further, the transfer of a patient to another physical therapist can be easily done.

### System-related factors (Outer setting)

#### Necessity to change efficiency

Stakeholders concluded that time is an advantage, but also a disadvantage. For instance, digital care could save time when filling out questionnaires in advance, however there is more time needed for preparation. All participants were in accordance that the financial reimbursement for blended care needed to be clarified (e.g. time for preparation and for digital care, costs for licenses and systems). Stakeholders determined that health insurance companies needed to cover the costs for in-person and digital care. Therefore, the single blended care intervention needed to be specified and described well.

#### Necessity of clear structural conditions

Structural preconditions were e.g. legal requirements, proof of effectiveness, data protection and security. Stakeholders suggested certifications for each type of technology, which meet data protection guidelines. Additionally, physical therapists suggested educating patients regarding data protection and the security.

An (advanced) training for physical therapists should especially focus on digital communication, data protection issues and evidence-based digital health. Patients should especially be educated regarding the handling of technology.

#### Clear rules and roles before an implementation

Stakeholders concluded that important steps of the implementation process are the communication and promotion of blended care, dealing with resistance, training of physical therapists as specialists and well-prepared introduction of technologies.

Structural facilitators were seen in the COVID-19 pandemic and if patients were provided with digital devices. The competitive market, missing transparency, privacy issues and different understandings of blended care were considered as structural barriers. All facilitators and barriers regarding blended physical therapy are listed in Figure 2 (Suppl. Material 4, Figure 2).

## Discussion

This study investigated different perspectives of patients, physical therapists and stakeholders on blended physical therapy of patients with OA.

Overall, patients and physical therapists are skeptical about blended physical therapy, which can be seen in the results of both. For instance, there was low patient acceptance of digital care, patients and physical therapists expressed the importance of in-person care and the integration of in-person treatment at the beginning and the end of each therapy section. However, blended physical therapy is currently unknown for both patients and physical therapists. Since it fits into future care models, it is still crucial to acquainting patients and physical therapists with blended physical therapy. Therefore, it is important to listen carefully to the preconditions, facilitators and barriers, which have being mentioned by the patients and physical therapists.

The most stated facilitators of blended physical therapy according to all participants were the individualization of blended physical therapy, the user-friendliness of the technology, the COVID-19 pandemic, access to digital devices and a stable internet connection. Barriers of blended physical therapy included technical skills of patients and physical therapists, costs, as well as the society’s lack of knowledge and information regarding blended physical therapy interventions.

One major finding was that the acceptance of digital care is still quite low in patients, whereas physical therapists are more open to use it. Interestingly, the Dutch *e-Exercise* project revealed this finding reversely (6). Patients were more enthusiastic and physical therapists more critical (6). This is quite remarkable, since it is probably due to the reason that the patients had experiences with a specific blended intervention, which obviously influenced their opinion and attitude towards blended physical therapy. Therefore, it seems crucial to gain positive experiences with blended physical therapy (23). In contrast, physical therapists had mixed experiences with *e-Exercise*, since e.g. the workload increased and it was more time-consuming, especially at the beginning (24). Patients, who did not have experience with digital care at all, were more skeptical and expected more barriers of it. A further personal precondition is the learning of new competences. Patients, as well as physical therapists seem to be open and willing to learn new competences, which can possibly increase the acceptance and change their attitudes regarding blended physical therapy (25, 26). This is also mentioned in previous studies as a key facilitator for the uptake and acceptance of digital care (25, 26).

An intervention-related precondition is to have a first and last in-person physical therapy session. This aspect was crucial for physical therapists, since they have difficulties to imagine performing a thorough first assessment or evaluation digitally (23, 27).

A further intervention-related precondition is the individualization of care. A key finding is that there is no “one-fits-all” solution, but rather the necessity to tailor blended physical therapy to the specific needs of each patient. This is mentioned as a main advantage of blended physical therapy, since it is beyond the borders of traditional care to provide, for instance, immediate and automated feedback specifically tailored to the patient (8, 23, 25). While they still have the opportunity to see their patient in-person and then have more time e.g. for in-depth conversations and personal attention. In general, physical therapists need to have the possibility to act flexible and to have the competence to decide, if a patient is suitable for blended care or not. The Dutch *Blended Physiotherapy Checklist* already supports and guides physical therapists in their clinical reasoning process while setting up a personalized blended physical therapy intervention (11).

Important preconditions regarding organizational factors are the interoperability of different software. Especially the physical therapists need to use different systems (e.g. administration, training programs) and without data transfer between the systems, it is not attractive to use such systems at all (28). Therefore, IT companies are responsible to develop interfaces between systems to enable interoperability. A change of facilities is also necessary in order to create enough privacy and a safe space for the physical therapist and the patient e.g. while having a video conference (23, 29).

The main system-related precondition is the reimbursement of blended physical therapy, which is also an issue in different countries (12, 29-31). Even though the COVID-19 pandemic enabled reimbursement of telehealth services, there is still no permanent solution (30). Since there is so far a lack of a payment solution, it is recommended to conduct pilot studies in order to investigate the usability and effectiveness of specific blended physical therapy approaches, having the mentioned preconditions, facilitators and barriers of this paper in mind. Furthermore, it is important to get a clear picture of data protection and safety issues. Stakeholders consent to have certificates for software, which help to get an overview as a user and rates technologies regarding their value, which is already existing in some countries (12, 29). Independent, public institutions might generate those guidelines, certificates and overviews for users. A further important system-related precondition was the development of an advanced training program for digital competences, which can be integrated in the curriculum of undergraduate and postgraduate physical therapist training programs. Therefore, it is necessary to create a framework of digital competences (32).

An important strength of this study is the investigation of blended physical therapy and not solely digital care. Simultaneously, it is challenging to investigate those two concepts separately, since they are very connected and participants had difficulties to distinguish between them. Therefore, parts of the results relate to digital care in general, not solely to blended physical therapy. A further strength is the inclusion of both the patient and the physical therapist’s perspective, which is complemented by a final discussion of stakeholders.

Additionally, the recruitment of two different groups of patients and physical therapists (with and without experience in digital health) contributed to a holistic picture. Limitations of our study are that our findings cannot be generalized to every type of blended physical therapy, since they may differ. Furthermore, two researchers held the interviews, which might have influenced the flow of the interviews in different ways. To prevent that, a topic guide was used, which supported covering the main topics.

Although, both patients and physical therapists were not too enthusiastic about blended physical therapy, consensus on needs and preconditions of blended physical therapy serves as a principal foundation for relevant caregivers, stakeholders and researchers. Needs, preconditions, facilitators and barriers have been indicated in four domains. The findings underline the importance to develop blended physical therapy interventions with a whole group of different stakeholders, which is crucial to facilitate the use and implementation of blended physical therapy at a later stage.

## Data Availability

Data are available on reasonable request. Data are in form of digital voice recording of interviews, which were also transcribed verbatim into Word files. Data of the online survey and demographics are available as excel files. Data of the focus groups are available in form of written protocols as Word files. These data are stored in a password-secured research drive, which is only accessible to Franziska Weber. Voice recordings contain identifiable data, which will not be available on request to maintain anonymity of the participants. The other files with de-identified participant data may be made available on reasonable request.

## Abbreviations

OA: Osteoarthritis

## Author Contributions

Concept/idea/research design: Franziska Weber, Corelien Kloek, Christian Grüneberg and Cindy Veenhof

Writing: Franziska Weber, Corelien Kloek, Angela Arntz, Christian Grüneberg and Cindy Veenhof

Data collection: Franziska Weber and Angela Arntz

Data analysis: Franziska Weber, Angela Arntz and Christian Grüneberg

Project management: Franziska Weber and Christian Grüneberg

Providing facilities/equipment: Christian Grüneberg

Providing institutional liaisons: Christian Grüneberg

Consultation (including review of manuscript before submitting): Franziska Weber, Corelien Kloek, Angela Arntz, Christian Grüneberg and Cindy Veenhof

## Acknowledgements

The authors thank all participants of the study for their generous participation.

## Ethics Approval

The Ethics Committee of the University of Applied Health Sciences in Bochum approved this study (201116_Grüneberg, 04.01.2021).

All participants gave written informed consent before data collection began.

## Disclosure and Presentations

The authors declare that there is no conflict of interest.

