## Supplementary Material 1-3 for "Blended care in patients with knee and/or hip osteoarthritis in physical therapy: a Delphi study on needs and preconditions"

### Supplementary Material 1: Semi-structured interview guides for the interviews with (1) the patients and with (2) the physical therapists

#### 1. Interview guide for patients

Guide for patients **who have experience with digital (health) care** and patients <u>who have no experience with digital (health) care</u> and questions for both groups.

**Research question:**

"What are the needs, preconditions, barriers and facilitators regarding blended physiotherapy in patients with knee and/or hip osteoarthritis from the perspective of patients, physiotherapists and further stakeholders of the health care system?"

Introduction (not recorded)

- Greetings and appreciation for taking time
- Brief description of the interview procedure and approximate duration, information on recording equipment
- Emphasis on the voluntary nature of the participation and the possibility of stopping the interview at any time
- Encouraging open and free narration, no evaluation of what is said and no right or wrong
- Clarification of questions

| No | Interview question | Questions to keep up the narrative flow | Operationalization |
|---|---|---|---|
| **Previous experience with physical therapy regarding osteoarthritis** | | | |
| **1** | What were the main aspects of your previous physiotherapeutic care regarding your osteoarthritis related symptoms? | • For instance, aspects are examination, information/education, exercises, physical activities and evaluation. | Characterization of the participants |
| **2** | What were important aspects of your previous physiotherapeutic care regarding your osteoarthritis related symptoms? | • Think about the aspects just mentioned and name those, which you consider as important. | Characterization of the participants |
| **Use and interaction with digital (health) applications** | | | |
| **3** | **How do you use digital (health) applications in your daily life?**<br>***(Definition of a digital (health) application)*** | • **Which digital (health) application do you use?**<br>• **For which purpose do you use digital (health) applications?**<br>• **Which device (PC, laptop, tablet, smartphone) do you use for your digital (health) applications?**<br>• **How often do you use digital (health) applications?** | Characterization of the participants |

| | | | |
|---|---|---|---|
| | | • **To what extent has the use of digital (health) applications in your daily life changed due to the Covid-19 pandemic?** | |
| **4** | Why are you not using digital (health) applications in your daily life?<br>*(Definition of a digital (health) application)* | • What is the reason for the non-usage?<br>• Please describe it in more detail. | Characterization of the participants |
| **5** | How would you rate your own skills in using digital (health) applications? | • Please specify.<br>• Please give examples. | Characterization of the participants |
| **Use of digital (health) care in general** | | | |
| **6** | What is your opinion on the use of digital (health) applications in health care?<br>What are the advantages and disadvantages? | • To which extent has, the Covid-19 pandemic changed your attitude towards digital (health) care.<br>• Please describe it in more detail.<br>• Please give an example to illustrate it in more detail. | Individual |
| **7** | What experience did you make within the health care sector regarding the use of digital (health) care? | • How did you experience it?<br>• Please give an example, so I can easily imagine, what you mean. | Individual |
| **Use of digital (health) care in physiotherapy** | | | |
| **8** | How do you imagine the ideal physiotherapeutic care in 10 years? | • Does your imagination change when you think of the possibilities of digitalization?<br>• Do you have any other suggestions? | Individual |
| **In between: presentation of a video as an example of a blended care situation: “Now, an explanatory video follows, providing an example of how a blended care approach can be performed in practice. At the end of the video, a written definition of blended care is given.”** | | | |
| **9** | What do you think about it, when you see this example of a blended care intervention? | • What are your first thoughts regarding this example? | Individual |
| **10** | What are barriers to implement blended care in physical therapy? | • Please describe it in more detail.<br>• Can you think of anything else that comes to your mind? | Individual |
| **11** | What are facilitators to implement blended care in physical therapy? | • Please describe it in more detail.<br>• Can you think of anything else that comes to your mind? | Individual |
| **12** | How would blended care change the physical therapist-patient-relationship? | • Can you think of anything else that comes to your mind? | Inner Setting |

| | | | |
|---|---|---|---|
| | | • Please describe it in more detail. | |
| **12** | Which skills/knowledge should you provide as a patient in order to implement blended care? | • Can you think of anything else that comes to your mind?<br>• Please describe it in more detail. | Inner Setting |
| **13** | Which conditions would have to be fulfilled to make blended care feasible in physical therapy? | • Think of technical, practical, personnel, financial and organisational (pre-) conditions.<br>• Can you think of anything else you would like to add? | Outer Setting |
| **14** | What are your needs for the content of digital (health) applications, embedded in a blended care approach regarding osteoarthritis? | • You mentioned earlier, that …. What do you mean with that?<br>• Can you think of anything else that comes to your mind?<br>• Please describe it in more detail. | Intervention |
| **15** | What treatment aspects/parts (examination, information/education, exercises, physical activities, evaluation) do you think would be suitable for online or in-person therapy? | • Which medium (video, video chat, app, pc program …) would be best suited for which aspect/part?<br>• Which aspect/part of the physiotherapeutic treatment should be delivered via which digital device (tablet, smartphone, pc, laptop …)?<br>• Why? | Intervention |
| **16** | What is the ideal percentage distribution of online or personal contact over the entire process of physical therapy for you? | • Please explain your opinion. | Intervention |
| **17** | Do you have any further comments on this topic? | • Are there any further questions?<br>• Is there anything, you would like to highlight? | Closure of the interview |

Finishing the interview (not recorded)

- Appreciation for the cooperation and small gift
- Note: contact details of the university and contact person for any open/further questions or for clarifying discussions

Definition: Digital (health) application

Digital (health) applications are programs/software that fulfil certain functions and are offered digitally (via smartphone, tablet, pc or laptop). Such as an app, that counts your steps.

#### 2. Interview guide for physical therapists

Guide for physical therapists **who have experience with digital (health) care** and physiotherapists <u>who have no experience with digital (health) care</u> and questions for both groups.

**Research question:**

"What are the needs, preconditions, barriers and facilitators regarding blended physical therapy in patients with knee and/or hip osteoarthritis from the perspective of patients, physical therapists and further stakeholders of the health care system?"

Introduction (not recorded)

- Greetings and appreciation for taking time
- Brief description of the interview procedure and approximate duration, information on recording equipment
- Emphasis on the voluntary nature of the participation and the possibility of stopping the interview at any time
- Encouraging open and free narration, no evaluation of what is said and no right or wrong
- Clarification of questions

| No | Interview question | Questions to keep up the narrative flow | Operationalization |
|---|---|---|---|
| Previous experience with physiotherapeutic care of people with osteoarthritis | | | |
| 1 | What are the **different aspects** regarding your physiotherapeutic care of patients with osteoarthritis? | • For instance, aspects are examination, information/education, exercises, physical activities and evaluation. | Characterization of the participants |
| 2 | What are **important aspects** regarding your physiotherapeutic care of people with osteoarthritis in your opinion? | • Think about the aspects just mentioned and name those, which you consider as important. | Characterization of the participants |
| 3 | To which extent does the treatment vary in relation to the status of osteoarthritis of your patients? | • Please explain. | Characterization of the participants |
| Use and interaction with digital (health) applications | | | |
| 4 | **How do you use digital (health) applications in your daily life?** | • **Which digital (health) application do you use?** | Characterization of the participants |

| | | | |
|---|---|---|---|
| | ***(Definition of a digital (health) application)*** | • **For which purpose do you use digital (health) applications?**<br>• **Which device (PC, laptop, tablet, smartphone) do you use for your digital (health) applications?**<br>• **How often do you use digital (health) applications?**<br>• **To what extent has the use of digital (health) applications in your daily life changed due to the Covid-19 pandemic?** | |
| 5 | <u>Why are you not using digital (health) applications in your daily life?</u><br><br><u>*(Definition of a digital (health) application)*</u> | • <u>What is the reason for the non-usage?</u><br>• <u>Please describe it in more detail.</u> | Characterization of the participants |
| 6 | How would you rate your own skills in using digital (health) applications? | • Please specify.<br>• Please give examples. | Characterization of the participants |
| Use of digital (health) care in general | | | |
| 7 | What is your opinion on the use of digital (health) applications in health care?<br><br>What are the advantages and disadvantages? | • To which extent has, the Covid-19 pandemic changed your attitude towards digital (health) care.<br>• Please describe it in more detail.<br>• Please give an example to illustrate it in more detail. | Individual |
| 8 | What experience did you make within the health care sector regarding the use of digital (health) care? | • How did you experience it?<br>• Please give an example, so I can easily imagine, what you mean. | Individual |
| Use of digital (health) applications in physical therapy | | | |
| 9 | How do you imagine the ideal physiotherapeutic care in 10 years? | • Does your imagination change when you think of the possibilities of digitalization? | Individual |

| | | | |
|---|---|---|---|
| | | • Do you have any other suggestions? | |
| In between: presentation of a video as an example of a blended care situation: "Now, an explanatory video follows, providing an example of how a blended care approach can be performed in practice. At the end of the video, a written definition of blended care is given." | | | |
| 9 | What do you think about it, when you see this example of a blended care intervention? | • What are your first thoughts regarding this example? | Individual |
| 10 | What are barriers to implement blended care in physical therapy? | • Please describe it in more detail.<br>• Can you think of anything else that comes to your mind? | Individual |
| 11 | What are facilitators to implement blended care in physical therapy? | • Please describe it in more detail.<br>• Can you think of anything else that comes to your mind? | Individual |
| 12 | To which extent would blended care change the physical therapist-patient-relationship? | • Can you think of anything else that comes to your mind?<br>• Please describe it in more detail. | Inner Setting |
| 13 | Which skills should you provide as a physical therapist in order to implement blended care? | • Can you think of anything else that comes to your mind?<br>• Please describe it in more detail. | Inner Setting |
| 14 | Which conditions would have to be fulfilled to make blended care feasible in physical therapy? | • Think of technical, practical, personnel, financial and organisational (pre-) conditions.<br>• Can you think of anything else, you would like to add? | Outer Setting |
| 15 | What are your needs for the content of digital (health) applications, embedded in a blended care approach regarding osteoarthritis? | • You mentioned earlier, that …. What do you mean with that?<br>• Can you think of anything else that comes to your mind?<br>• Please describe it in more detail. | Intervention |

| | | | |
|---|---|---|---|
| 16 | What treatment aspects/parts (examination, information/education, exercises, physical activities, evaluation) do you think would be suitable for online or in-person therapy? | • Which **medium** (video, video chat, app, pc program …) would be best suited for which aspect/part?<br>• Which aspect/part of the physiotherapeutic treatment should be delivered via which **digital device** (tablet, smartphone, pc, laptop …)?<br>• Why? | Intervention |
| 17 | What is the ideal percentage distribution of online or personal contact over the entire process of physical therapy for you? | • Please explain your opinion. | Intervention |
| 18 | Do you have any further comments on this topic? | • Are there any further questions?<br>• Is there anything, you would like to highlight? | Closure of the interview |

#### Finishing the interview (not recorded)

- Appreciation for the cooperation and small gift
- Note: contact details of the university and contact person for any open/further questions or for clarifying discussions

#### Definition: Digital (health) application

Digital (health) applications are programs/software that fulfil certain functions and are offered digitally (via smartphone, tablet, pc or laptop). Such as an app, that counts your steps.

#### Supplementary Material 2: List of themes and subthemes of the data analysis of the interviews

| List of Themes | Memo |
| --- | --- |
| Themes and subthemes | |
| **Implementation process** | |
| **Patient-related factors** | This code is used when it concerns the factors, which influence the use/usage of blended care. |
| Role concept | This code is used when talking about how the patient understands his or her role and the physiotherapists role in the treatment process and what influence the blended care concept has on this understanding of the role and what preconditions must be fulfilled for this change to take place. |
| Relationship | This code is used when talking about the changes in the relationship between the physiotherapist and the patient from the patient's point of view due to the use of blended care and when stating which conditions must be fulfilled so the relationship does not change negatively despite the use of blended care. |
| Competences | This code is used to indicate which competences the patient needs to have at a personal level in order to use blended care. |
| **Acceptance** | |
| Role of physical therapy | |
| Digital tools | |
| **System-related factors** | |
| Time | This code is used when considering whether the implementation of blended care gives the user more or less time for the treatment process. |
| Costs/ expenses | This code is used to identify the costs of implementing blended care and who or what system is responsible for covering the costs. Furthermore, it is also included if it is a precondition for the implementation of blended care that the costs are covered. |
| Data security and patient rights | This code is used if subjects talk about privacy within blended care and if factors are named that require the patient's consent. |
| Structural (pre-)conditions/ framework conditions | Code is used when talking about changes in work, therapy and acceptance of digital media due to the pandemic and when talking about preconditions for use and structural preconditions for implementing blended care at a system level. |
| **Intervention-related factors** | |
| Digital elements | |

| | |
|---|---|
| Quality | This code is used when talking about factors that influence the implementation of blended care and when subjects describe how the quality of care changes because of blended care. |
| Allocation of the competences | |
| Order | |
| **Technical factors** | This code is used when it generally concerns technical requirements for implementation of blended care. |
| Software | This code is used when talking about formats (video, text form, video chat, etc.), which content should be offered, the functional structure of the online program and which functions it should have. It is also used when talking about the need for WLAN access in order to be able to use blended care. |
| Hardware | This code is used when it comes to access to digital devices and when it comes to the allocation of digital components to the end devices. |
| Usability | This code is used when it comes to the requirements that must be fulfilled in order to use digital tools within the blended care intervention and to simplify the usability of the technology. |
| **Organisational factors** | |
| Practical setting | |
| Cooperation/ collaboration | |
| Working conditions | |
| Personal structure | |
| **Therapist-related factors** | |
| Treatment approach | This code is used when the participant talks about, how he/she is currently treating his/her patients and when he/she talks about how his/her therapy is changing due to the use of blended care. |
| Relationship | This code is used when talking about changes in the relationship between therapists and patients from the therapist's point of view due to the use of blended care and when stating which conditions have to be fulfilled so that the relationship does not change negatively despite the use of blended care. |
| Competences | This code is used when talking about which competences the therapist requires on a personal level in order to apply blended care. |

#### Supplementary Material 3

Table 5 Preferred mode of the therapy component: online, in-person or online and/or in-person out of the perspective of physical therapists and patients (n=16)

| **Therapy components** | **Patients (n=6)** | **Physical therapists (n=9)** |
|---|---|---|
| First therapy session/ getting to know | In-person (n=6) | In-person (n=5) |
| Information/ education session | In-person (n=5) | Online and/or in-person (n=5) |
| Consultation | Online and/or in-person (n=4) | Online and/or in-person (n=7) |
| Screening process/ diagnostic process | In-person (n=4) | Face-to-face (n=6) |
| Instruction of exercises | In-person (n=4) | Online and/or in-person (n=5) |
| Functional integration of movement into activities of daily living | In-person (n=4) | Online and/or in-person (n=8) |
| Evaluation/ last therapy session | In-person (n=6) | Online and/or in-person (n=9) |

Table 6 Preferred ratio of online and in-person therapy of patients with osteoarthritis

| **Ratio in percentages** | **Patients (n=7)** | **Physical therapists (n=9)** |
|---|---|---|
| 0% online / 100% in-person | **1** | 0 |
| 10% online / 90% in-person | 0 | 0 |
| 20% online / 80% in-person | **1** | 0 |
| 30% online / 70% in-person | **1** | 0 |
| 40% online / 60% in-person | **1** | 0 |
| 50% online / 50% in-person | **1** | **2** |
| 60% online / 40% in-person | **1** | **3** |
| 70% online / 30% in-person | **1** | **4** |
| 80% online / 20% in-person | 0 | 0 |
| 90% online / 10% in-person | 0 | 0 |
| 100% online / 0% in-person | 0 | 0 |

Table 7 Checklist of practice setting regarding blended physical therapy in the perspective of patients and physical therapists

| **Practice setting checklist** | |
|---|---|
| Patients | Physical therapists |
| Separate rooms (enough space, calm, privacy) | Facilities (enough space, privacy) |
| Home office physiotherapists (privacy) | Possibility and preparation of home office for physical therapists |
| Equipment must be available | |
| Technology (devices and WLAN) | Technology (devices and WLAN) |
| | Data protection |
| | Concepts should be clear in advance |
| | Proper time schedule |

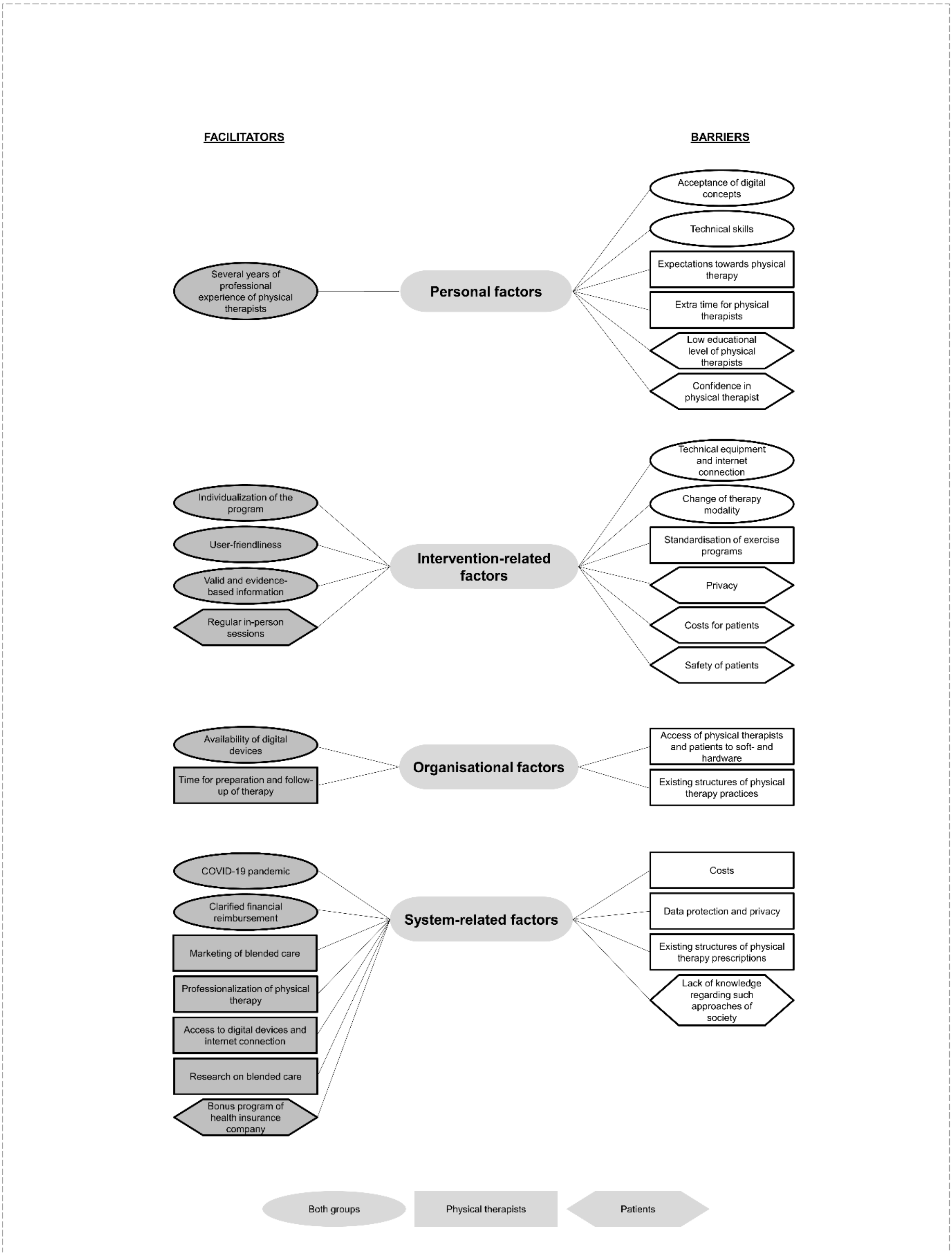


*Figure 1 Facilitators and barriers of blended physical therapy*
